# COVID-19 Vaccine Uptake And Its’ Associated Factors among general population In Basmaia City in Baghdad 2022

**DOI:** 10.1101/2023.04.07.23288262

**Authors:** Hussein Abdalrahim Saber, Mohammed Asaad Albayaty

## Abstract

**Objective:** Vaccination is a vital cornerstone of public health, which has saved countless lives throughout history. Therefore, achieving high vaccination uptake rates is essential for successful vaccination programs. Unfortunately, vaccine uptake has been hindered by deferent factors and challenges. The objective of this study is to assess COVID-19 vaccine uptake and associated factors among the general population.

**Methods:** This study is a descriptive cross-sectional study conducted in Basmaia city, Baghdad from June to October 2022. Data were collected through a semi-structured questionnaire using multi-stage random sampling. Statistical analysis was performed using descriptive statistics, chi-square analysis, Mann-Whitney test, and binary and multivariable logistic regression.

**Results:** The prevalence of COVID-19 vaccine uptake was 70.4%. The most common reason for getting vaccinated was protection from the disease, while fear of side effects and not needing the vaccine were the main reasons for refusal.

The study found that gender, age, education level, job title, risk perception, knowledge, and attitude towards the vaccine were significantly associated with COVID-19 vaccine uptake. Males were 2.273 times more likely to get vaccinated than females, and older age groups had higher odds of vaccination than younger age groups. Those with higher education levels were also more likely to receive the vaccine. Participants with higher risk perception, knowledge, and positive attitude towards the vaccine were more likely to get vaccinated.

And found that mandatory vaccination policies may negatively impact uptake of subsequent vaccine doses.

**Conclusion:** The study found a high prevalence of COVID-19 vaccine uptake, with gender, age, education level, and job title being significant factors associated with vaccine uptake. Additionally, mandatory vaccination policies may have a negative impact on the uptake of subsequent vaccine doses. Public health efforts should prioritize addressing these factors to increase vaccine uptake.

## Introduction

The COVID-19 outbreak started in Wuhan, China in December 2019 and rapidly spread to other countries, leading the World Health Organization to declare it a global pandemic.(1)(2) As of September 2022, there have been 612 million confirmed cases and 6.5 million deaths worldwide.(3) In response, the World Health Organization (WHO) led a global campaign for the prevention, early diagnosis, and treatment of the disease.(4) The development of a vaccine was seen as the definitive solution to end the crisis,(5) and multiple vaccine candidates were developed and tested globally.(6) Four vaccines were authorized for use in multiple countries, including mRNA-based vaccines, an adenovirus vector platform, and inactivated virus technology.(7)(8)

Vaccination is crucial in preventing the spread of infectious diseases and mitigating associated morbidity and mortality.(9) In the context of the COVID-19 pandemic, high vaccination rates are necessary to provide indirect protection, return to normal societal patterns, reopen the economy,(10) and achieve herd immunity.(11) However, there is a challenge of low vaccine uptake due to concerns about the safety and efficacy of new vaccines.(12)

In Iraq, the COVID-19 pandemic has reached alarming proportions with 2.5 million confirmed cases and 25,000 deaths as of September 2022.(13) The government has made efforts to provide access to vaccines for its citizens at no cost and has implemented a vaccination card strategy to incentivize vaccination which requires individuals to present a valid vaccination card for entry into government buildings, shopping centers, educational institutions, gyms, and other places.(14) Despite these efforts, vaccine coverage remains low with only 18% of the population having received full doses as of September 2022.(15) Addressing vaccine low coverage is a major challenge to achieve herd immunity and effectively combat the pandemic.

Vaccine uptake can be defined as the number of people vaccinated with a certain dose of the vaccine when it is made available to them in a certain time period, which can be expressed as an absolute number or as the proportion of a target population. This metric is a crucial determinant of a population’s immunity to a particular disease and provides insight into the overall efficacy of a vaccine program.(16)

There are three main methods for measuring vaccine uptake: Routine Monitoring(direct measurement), Periodic Surveys, and Indirect Measurements.

Routine Monitoring involves collecting data from health facilities and providers to track vaccine uptake. Data is collected through various sources such as Health Management Information Systems (HMIS), Electronic Immunization Registries (EIR), and electronic Vaccination Cards. This method provides a comprehensive view of vaccine uptake as it captures data from all sources of vaccine administration.

Periodic Surveys involve conducting surveys to assess vaccine uptake and service delivery. The surveys can be either population-based or facility-based, which provide a snapshot of the current state of vaccine uptake. They can be used to track changes over time and make evidence-based decisions to improve vaccine programs and increase vaccine uptake.

Indirect Measurements use models to estimate vaccine uptake based on available data such as disease incidence or seroprevalence data. This method is useful for estimating vaccine uptake in populations where direct measurement is not possible, but it is less accurate.(16)

Prior to the COVID-19 pandemic, Low vaccination uptake has been observed in numerous disease outbreaks such as dengue(17), malaria(18), Ebola(19), chikungunya(20), and monkeypox(21). and is a persistent challenge for public health initiatives aimed at improving vaccine uptake.

The prevalence of low vaccination uptake can be attributed to various factors, including a lack of adequate knowledge regarding vaccines, unfavorable attitudes towards vaccines, and the proliferation of false information through traditional and social media, such as conspiracy theories. Additionally, concerns about vaccine efficacy and safety, including worries about both short and long-term side effects, have also been cited as reasons for low uptake. Other factors that can impact vaccination uptake include health conditions such as chronic diseases, risk perception of infection, religious beliefs, and political considerations.(22) Moreover, the COVID-19 pandemic has presented numerous challenges with regards to vaccine acceptance and uptake. Firstly, the magnitude of the vaccination effort is unparalleled as the aim is to offer the initial series of COVID-19 vaccines to the global adult population. In many regions, there are no established adult immunization programs, making it difficult to administer the vaccine even when doses are available. In areas where adult immunization programs exist, such as for influenza, uptake has often been low. Secondly, the COVID-19 information overload, which includes a mixture of accurate and inaccurate information, makes it difficult for individuals to access reliable information about vaccination. Misinformation and disinformation can significantly decrease vaccine acceptance(23) and may impact the advice given by healthcare providers.(24) Political decisions related to the pandemic have also been influenced by this information overload and political beliefs about the risk of COVID-19 and the effectiveness of preventive measures.(25) Finally, the prolonged duration of the pandemic has resulted in “pandemic fatigue”, leading to a decrease in motivation to follow public health recommendations, including vaccination.(26)

In conclusion, vaccine uptake is influenced by a complex interplay of factors, including risk perception, knowledge, previous exposure, demographic factors and attitudes towards vaccines. Understanding these factors and how they interact is essential for developing effective strategies to increase vaccine uptake and protect populations from preventable diseases. Accurate and timely measurement of vaccine uptake, along with an understanding of the factors that influence vaccine uptake, is critical for the success of vaccine programs.(26) And given the potential emergence of new COVID-19 variants at any time and the necessity of a new wave of mass vaccination, and the limited number of studies to date that have investigated COVID-19 vaccine uptake and its factors among the general population of Iraq, our study aimed to attain a more comprehensive understanding of the factors influencing COVID-19 vaccine uptake in Iraq. Our study specifically focused on sociodemographic factors, prior experience, risk perception, knowledge, attitude, and barriers to vaccination as they may impact vaccination decisions. This information is of utmost importance for government and policy makers to identify and address any barriers to vaccine distribution and to ensure a high rate of vaccine uptake among the population.

## Methodology

### Study design

A descriptive cross-sectional study was conducted in Basmaia city in Baghdad from June to October 2022 with the primary objective of assessing covid-19 vaccine uptake and its associated factors among the general population.

### Study population and Eligibility criteria

The study population included households whose members were 18 years or older, who resided in Basmaia city, and who were capable of reading and comprehending the Arabic language. Participants who agreed to take part in the study and completed the questionnaire were included in the analysis. However, incomplete responses were excluded from the analysis.

### Sample size

The sample size for this study was determined based on the following assumptions: a 95% confidence level, a margin of error of ±5%, and an estimated population size of 75,840 individuals living in the city.(27) The official vaccine uptake rate was estimated to range from 18% for two doses to 25% for one dose. Using the equation n = [z2 * p * (1 - p) / e2] / [1 + (z2 * p * (1 - p) / (e2 * N))], where z = 1.96, p = proportion of vaccine uptake (expressed as a decimal), N = population size, and e = margin of error, so the required calculated sample size was 260 participants. However, the study ended up collecting data from 310 individuals. Out of these, 70 participants had missing data, leaving a final sample size of 240 participants that was deemed suitable for analysis.

### Sampling method

The sampling method used in this study involved the selection of individuals from the general population of the city through multi-stage random sampling. The sampling process proceeded as follows (Figure 1):

**Figure 1.**
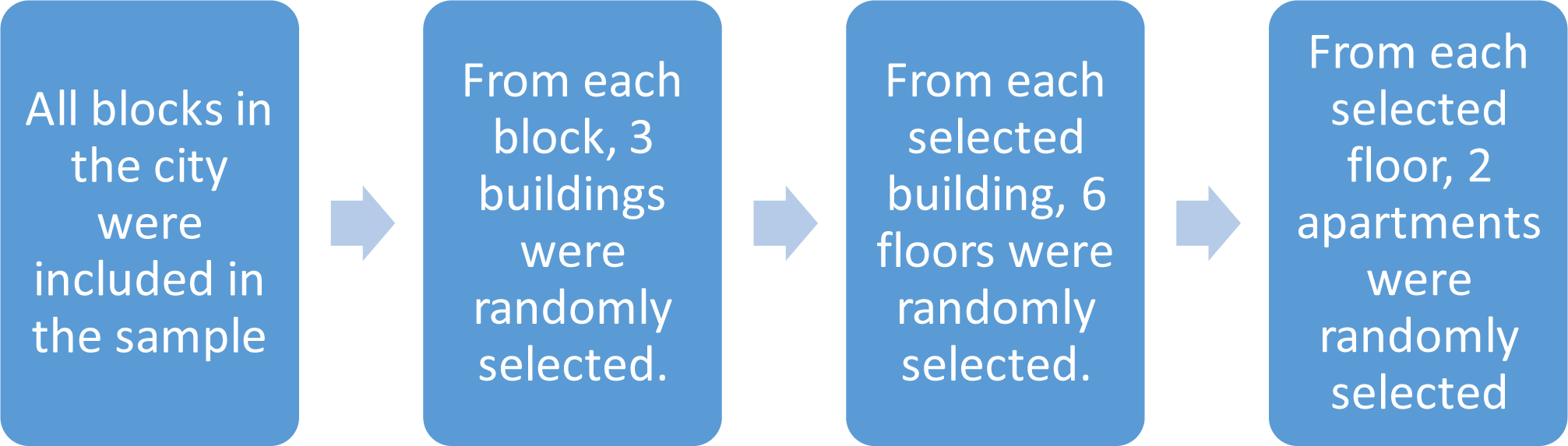
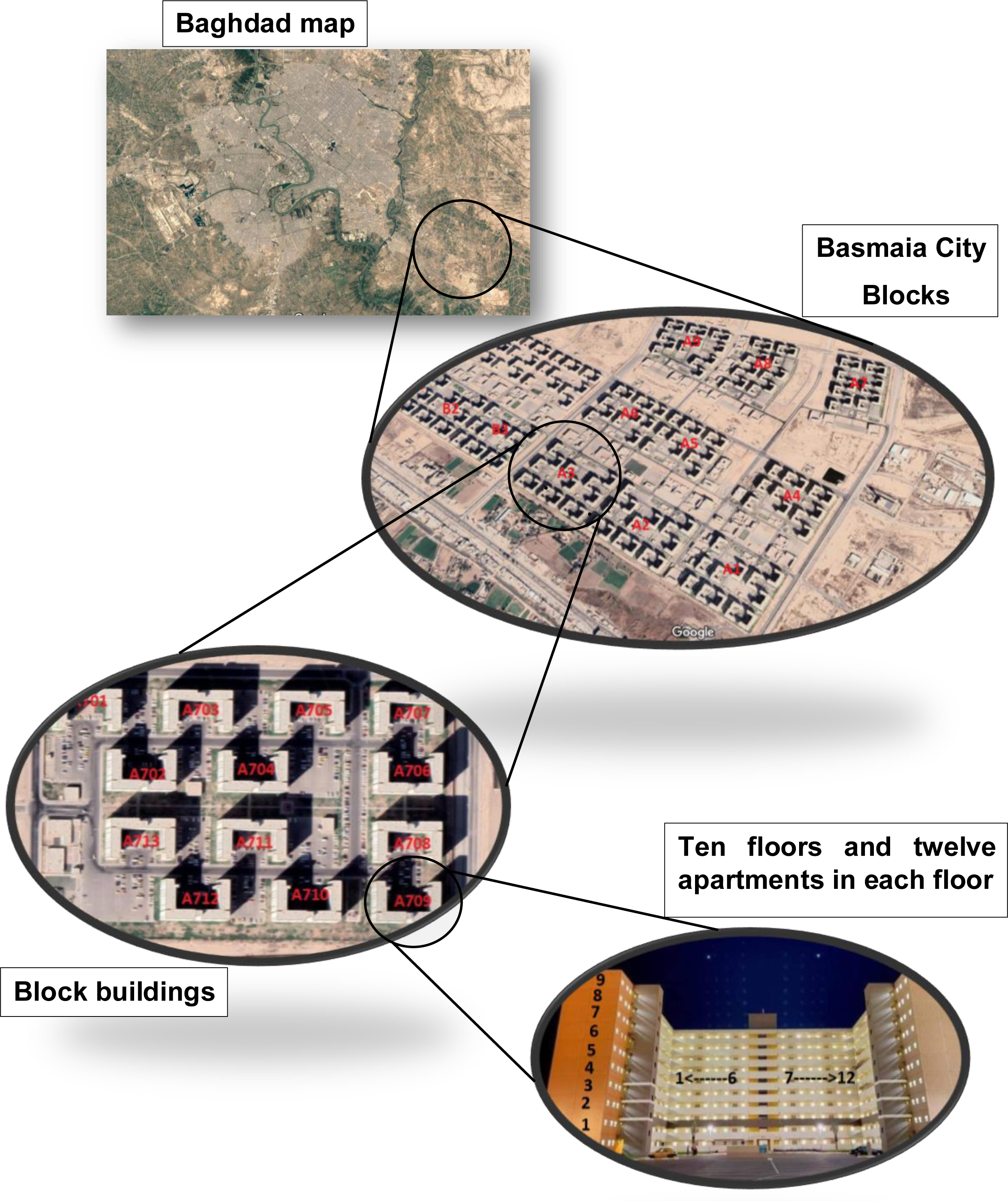

All the blocks in the city, which totaled to 12 (A1,A2-A9,B1,B2,B3), were included in the first stage.

In the second stage, three buildings from each block were selected using simple random sampling.

In the third stage, six floors from each selected building were randomly selected.

Finally, in the fourth stage, two apartments were randomly selected from each floor. To maintain the randomness of the sampling process, in cases of a vacant apartment, the apartment to the right was chosen.

The multi-stage random sampling technique used in this study was chosen to minimize any biases that could have occurred while selecting the sample, thus increasing the generalizability of the study findings to the target population.

### Data collection procedure

The data collection procedures were conducted from 3 pm to 7 pm one day in a week. during which time it was assumed that all household members would be at home. using the self-administered questionnaire, along with a pen, was distributed to all households with members aged 18 years or above, after obtaining informed consent, and were asked to complete it at their convenience. The researcher was available to provide assistance to participants who had questions or required clarification on any aspect of the questionnaire.

Upon completing the sampling of the building, the researcher revisited each apartment to retrieve the questionnaire with the pen and ensure that the data collection process was proceeding as planned. Furthermore, the researcher was available to answer any medical-related questions or concerns that the participants may have had.

Finally, at the end of the data collection procedure, a brochure containing updated information about COVID-19 vaccine facts was distributed to all participants to provide them with additional information. The distribution of the brochure was intended to promote awareness and education among the participants about the COVID-19 vaccine.

### Data collection instrument

Data were collected using a semi-structured questionnaire that was adapted by reviewing relevant literature and modified to suit the local context.(28)(29)(30)(31)(32)(33) To ensure the questionnaire’s reliability and validity, it was pre-validated by three independent reviewers and pretested on ten individuals to assess the clarity, comprehensiveness, sensitivity, and length of the questions. The questionnaire was first prepared in English and then translated into Arabic by a professional translator to ensure its comprehensibility to all participants. The questionnaire contained a brief introduction to the study’s background, objectives, eligibility criteria, voluntary nature of participation, and declaration of confidentiality and anonymity. Each participant took approximately six minutes to complete the questionnaire. The pilot study results were used to enhance the quality and efficiency of the primary survey, but they were not included in the final analysis of the study.

## Questionnaire measures

### Dependent variables

1. The main dependent variable of this study was the uptake of the COVID-19 vaccine. Vaccine uptake was defined as the proportion of participants who had received at least one dose of the COVID-19 vaccine, as determined by a closed-ended question asking, “Have you been vaccinated with the COVID-19 vaccine?”. Participants who answered “Yes” were classified as having received the vaccine, while those who answered “No” were classified as having not received the vaccine. (16)

2. Second dose uptake was also assessed among participants who reported receiving at least one dose of the COVID-19 vaccine.

### Independent variables

1. Socio-demographic and economic variables: The study included several independent variables related to socio-demographic and economic factors, including age, sex, academic achievement, job (health field, outside health field, other like no job, retired, housewife or student), and economic variables (family ownership of a house).

2. Chronic illness or comorbidities: Participants were asked if they had any chronic illness or comorbidities that may impact their risk perception and decision to receive the COVID-19 vaccine.

3. Experience with COVID-19: Participants were asked about their experience with COVID-19, including whether they had been previously infected, the perceived severity of their symptoms if infected (mild, moderate, severe), and whether any family member had suffered from a severe disease or death due to COVID-19.

4. Risk perception of getting COVID-19: Risk perception was assessed using a three-item questionnaire that asked participants to rate their concern about contracting COVID-19 at work, outside of work, and infecting their family or friends with COVID-19. Participants responded on a three-point Likert scale, with scores ranging from 3 to 9.

5. Knowledge about COVID-19 vaccine: the level of knowledge about the COVID-19 vaccine was assessed as an independent variable. A self-developed scale was used, which included seven items that were formulated based on the information presented by the official website of the World Health Organization.(34) The response categories for the knowledge items were “True”, “False” or “don’t know”. A correct answer was assigned 1 point, while an incorrect answer or “don’t know” was assigned 0 points. The scores of all the responses to the seven items were summed up to create one index, with a higher score indicating a higher level of knowledge. As the development of COVID-19 vaccines is still in progress, this approach is based on prior research’s approach to measuring knowledge about novel and emerging health-related issues.

6. Attitude towards COVID-19 vaccine: Participants’ attitudes towards the COVID-19 vaccine were assessed using a five-item questionnaire with five-point Likert-type responses. The total score ranged from 5 to 25, with higher scores indicating more positive attitudes towards the vaccine.

7. Reasons for uptake or refusal: Participants were asked to indicate their reasons for accepting or refusing the COVID-19 vaccine.

8. Mandatory uptake of the vaccine due to vaccine card strategy for entering governmental places or for travel and its impact on the uptake of subsequent vaccine doses.

## Data analysis

The statistical analysis for this study was performed using IBM SPSS Statistics version 26. Descriptive statistics, such as frequency distribution, percentage, and mean scores, were used to analyze the data, and the results were presented using tables, graphs, and charts. Chi-square analysis was conducted to explore the association between dependent and independent variables, and Mann-Whitney test was used for non-parametric continuous variables. Variables with p-values ≤ 0.05 on chi-square analysis and Mann-Whitney test were considered as potential candidates for binary logistic and multivariable logistic regression. These regression models were employed to investigate the relationship between the outcome variable and predictor variables. Independent variables with p-values <0.05 and adjusted odds ratios at a 95% confidence interval were identified as significant predictors of the outcome variable.

### Ethical consideration

This research study received approval from the Iraqi Board of Medical Specialization, with an approval number of 506. The need for consent for the acquisition, interpretation, and publication of the prospectively collected anonymous data for this research was waived by the Iraqi Board of Medical Specialization. Additionally, an official approval was obtained from the Basmaia City Council.

Before participating in the survey, individuals were given the opportunity to decline participation in the questionnaire. Participation in this study was entirely voluntary, with no coercion or compensation offered. Participants provided informed, verbal consent. Throughout the study, anonymity and confidentiality were maintained, with the data presented only in aggregated statistics. All data were saved in a secure file, with only the researchers in charge of the questionnaire having access to the information.

In consideration of misinformation, a brochure containing updated information about the COVID-19 vaccine was distributed to all households at the end of the questionnaire.

Furthermore, this study was conducted in accordance with the World Medical Association’s Declaration of Helsinki, which outlines ethical principles for medical research involving human subjects.

## Results

### Characteristics of the participants

The study involved 240 residents who completed the survey questionnaire. The majority of respondents (33.8%) were within the age range of 35-44 years, with a mean age of 37.5 and a standard deviation of 11.7. The study had an equal gender distribution1:1 ratio. The educational level of the participants showed that the majority of the respondents had a bachelor’s degree or institute degree, which accounted for 88 (36.7%) of the participants. This was followed by those with middle school degree (intermediate or secondary), with 41 (17.1%) participants. 165(68.8%) of participants reported having one or more chronic illness or comorbidities.155 (64.6%) reported owning a house.(Table-1)

**Table 1:** Socio-Demographic and Characteristics of the Study Participants Toward COVID-19 Vaccine Uptake and Associated Factors Among Basmaia city inhabitants 2022 (n=240)

| variables | Categories | Frequency (%) |
| --- | --- | --- |
| Age | 18-24 | 35(14.6%) |
|  | 25-34 | 60(25%) |
|  | 35-44 | 81(33.8%) |
|  | 45-54 | 42(17.5%) |
|  | >54 | 22(9.2%) |
| Gender | Male | 120(50%) |
|  | Female | 120(50%) |
| Education level | Primary school or below | 34(14.2%) |
|  | Middle school (intermediate or secondary) | 76(31.7%) |
|  | Bachelor or institute degree | 116(48.3%) |
|  | Master or PhD | 14(5.8%) |
| House ownership | Yes | 155(64.4%) |
|  | No | 85(35.4%) |
| Job title | Outside the health field | 143(59.6%) |
|  | Health field | 22(9.2%) |
|  | Other(no job, housewife, retired) | 75(31.3%) |
| Chronic illness or comorbidities | Have one or more chronic illness | 165(68.8%) |
|  | Do not have chronic illness | 75(31.3%) |

### The Prevalence of COVID-19 vaccine uptake

The prevalence of COVID-19 vaccine uptake among the general population in Basmaia City, Baghdad was 169 (70.4%). Of those who were vaccinated, the majority 146 out of 169 (86.4%) reported taking the second dose of the COVID-19 vaccine (60.8% of total). The most common reason reported for getting vaccinated was protection from the disease (54.6%), followed by being forced to vaccinate (10.8%) and receiving a recommendation from family (3.3%).(figure 3) On the other hand, the most frequently reported reasons for refusing the vaccine were fear of side effects (52.5%) and not needing the vaccine (37.2%). A small proportion of participants (5-8.4%) reported not being able to take the vaccine due to an underlying condition or not finding the vaccine they wanted.(figure 4)

**Figure 3.**
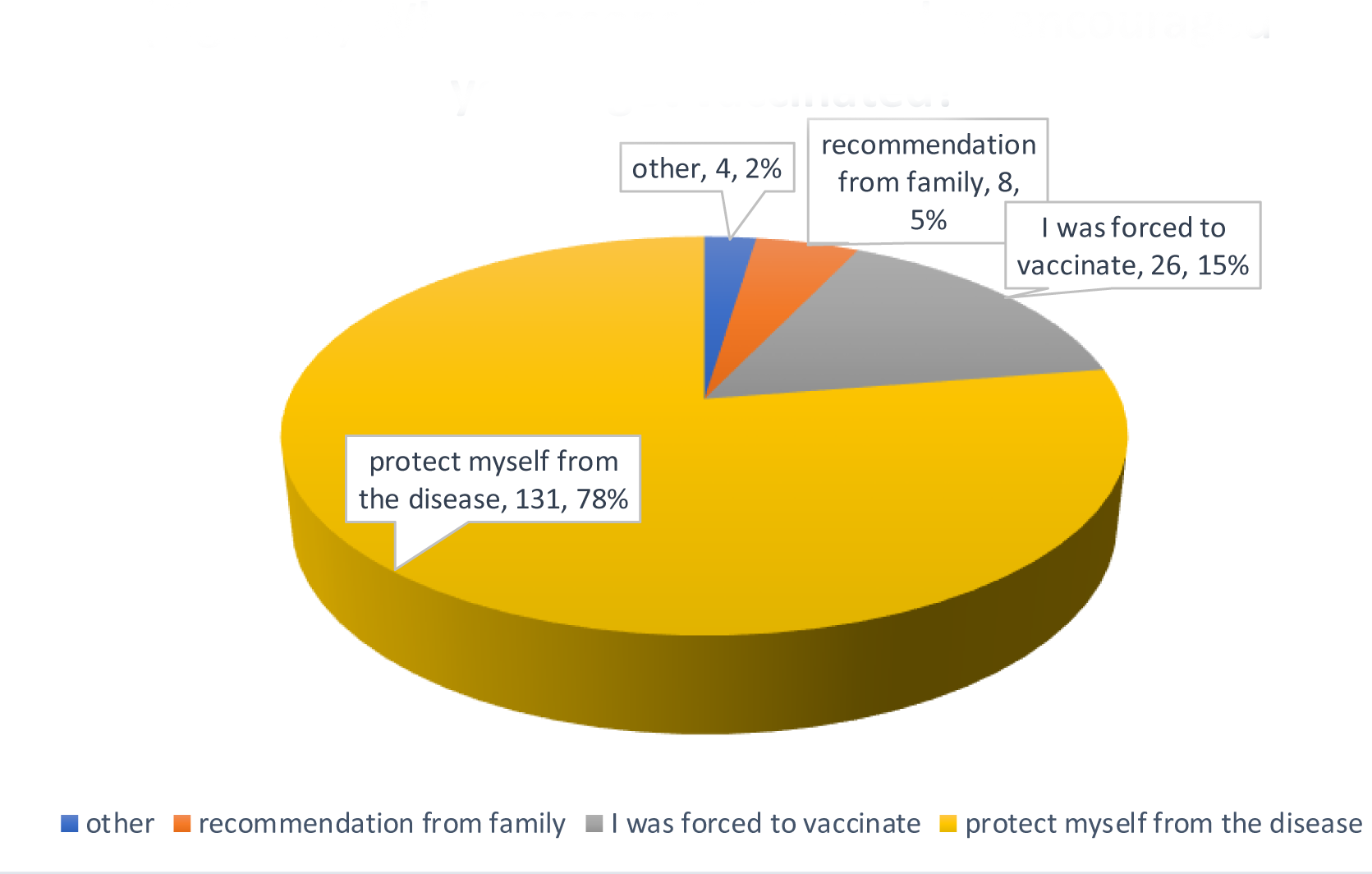
What reasons influenced or encouraged you to get vaccinated?

**Figure 4.**
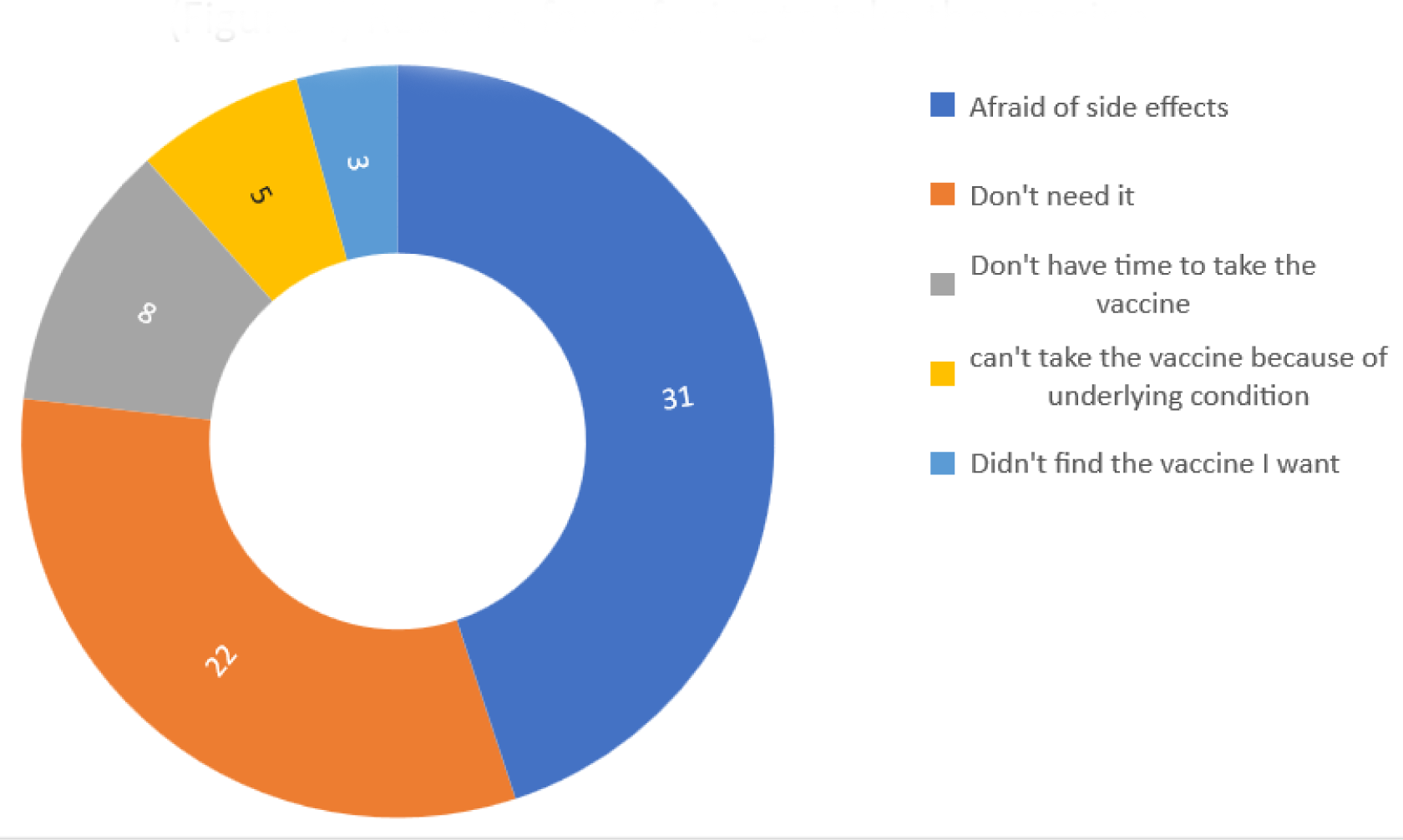
Reasons for refusing to take the vaccine

### Factors Associated with COVID-19 Vaccine Uptake

The chi-square analysis indicated that several variables, including gender, age, education level, and job title, were significantly associated with COVID-19 vaccine uptake (p < 0.05), as presented in (Table 2). However, some variables, such as house ownership, having chronic illness or comorbidities, previous infection, severity of symptoms, and death or severe illness in the family due to COVID-19 disease, were not statistically significant (p > 0.05) thus were excluded from binary and multivariate logistic regression analysis.

**Table 2:**
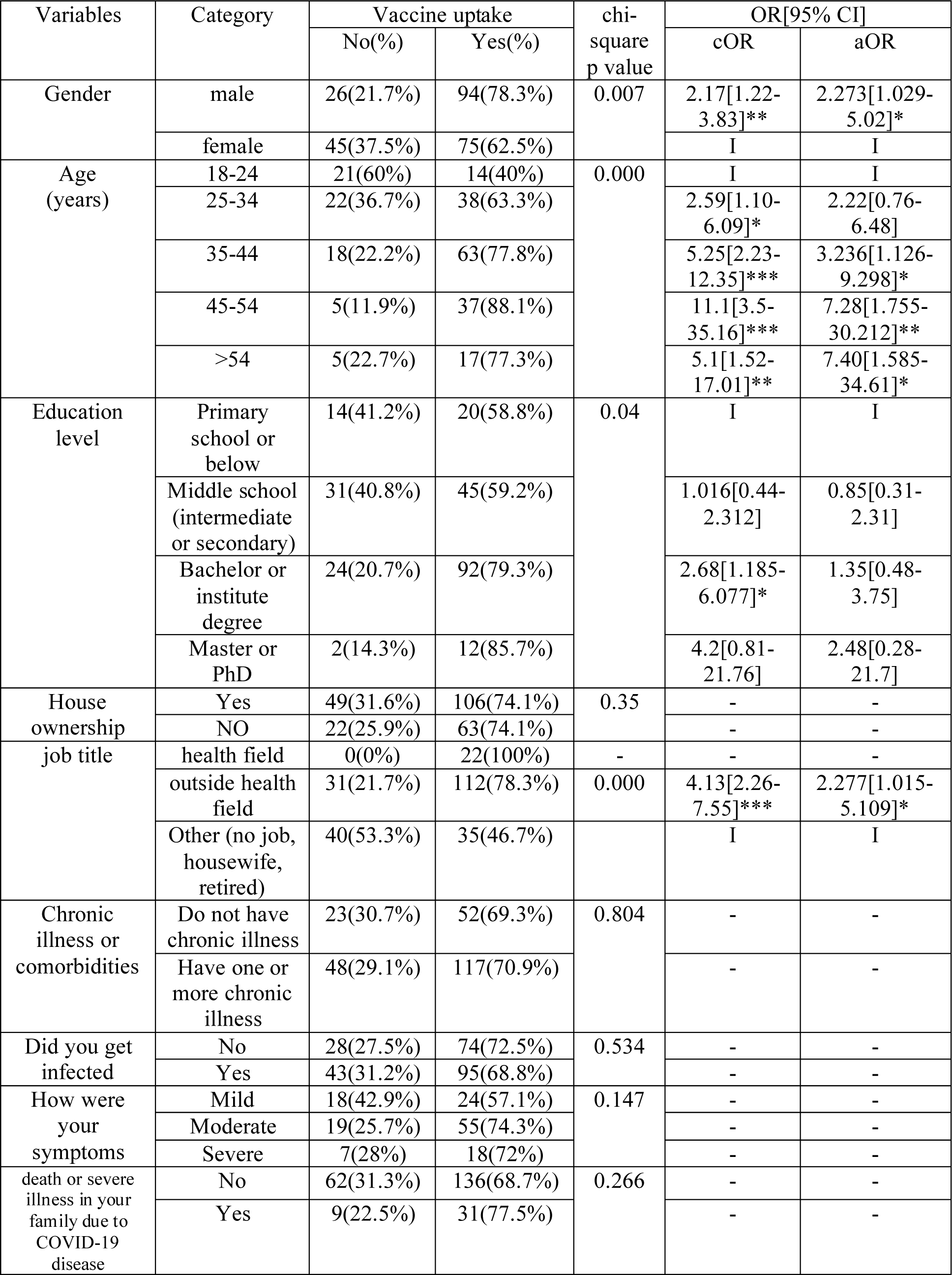
Association between demographic and health-related factors and COVID-19 vaccine uptake

The logistic regression analysis showed that males were significantly more likely to receive the COVID-19 vaccine compared to females, with a 2.273 times higher odds ratio (adjusted odd ratio) (95% CI [1.03-5.02]).

In terms of age groups, those aged 45-54 years and >54 years were found to have 7 times higher odds of getting vaccinated (aOR 7.28, 95% CI [1.7-30.2]; aOR 7.4, 95% CI [1.5-34.6], respectively) compared to the young age group (18-24 years). The 35-44 years age group was also significantly more likely to receive the vaccine, with a three times higher odds ratio (aOR 3.23, 95% CI [1.2-9.3]).

Participants with a higher education level (bachelor or institute degree) were more likely to receive the vaccine than those with a lower education level (primary school or below) (cOR=2.68,95% CI [1.18-6.07]).

Participants working in the health field had a 100% vaccine uptake rate, therefore logistic regression was not conducted for this variable. However, participants working outside the health field were more likely to receive the vaccine than those who were not employed or worked as housewives or retirees (OR=2.277, 95% CI [1.015-5.109]).

Among those who did not get infected with COVID-19, 74 out of 102 (72.5%) got vaccinated, compared to 95 out of 138 (68.8%) of those who did get infected. The difference in vaccine uptake between the two groups was not statistically significant (p=0.534). Participants with a family history of severe illness or death due to COVID-19 were more likely to receive the vaccine than those without such a history, although the association was not statistically significant.

There are significant associations between vaccine uptake and risk perception, knowledge, and attitude towards the vaccine (Table 3).

**Table 3:** Association between Vaccine Uptake and Risk Perception, Knowledge, and Attitude towards COVID-19 Vaccine

| Variables | Vaccine uptake<br>Mean Rank |  | Mann-<br>witney<br>test p<br>value | OR[95% CI] |  |
| --- | --- | --- | --- | --- | --- |
|  | NO | Yes |  | cOR | aOR |
| Risk perception<br>(3-9) | 97.71 | 130.07 | 0.001 | 1.378[1.153-1.64]*** | 1.258[1.001-1.58]* |
| knowledge (0-7) | 82.13 | 136.62 | 0.001 | 1.995[1.548-2.57]*** | 1.562[1.154-2.11]** |
| attitude (9-25) | 83.11 | 136.21 | 0.001 | 1.184[1.184-1.443]*** | 1.232[1.095-1.385]*** |

**Table 4:**
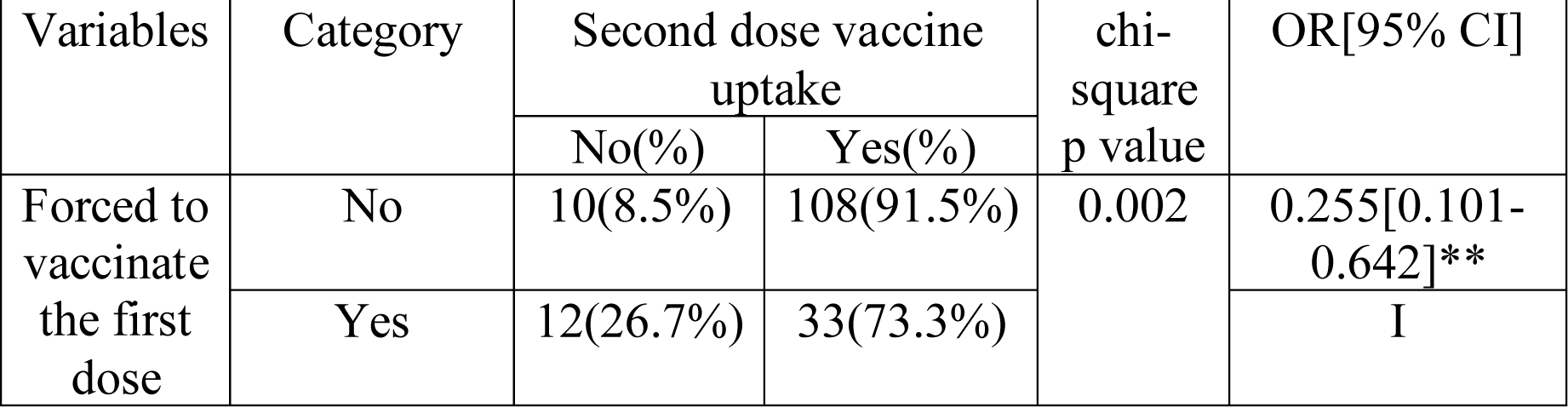
Second dose vaccine uptake by whether or not individuals were forced to vaccinate for the first dose.

The mean ranks of those who received the vaccine were higher for all three variables, indicating that those who were vaccinated tended to have higher scores for risk perception, knowledge, and attitude towards the vaccine. The Mann-Whitney test showed significant differences between those who received the vaccine and those who did not in terms of their risk perception (p=0.001), knowledge (p=0.001), and attitude (p=0.001).

The adjusted odds ratio for these variables suggest that after adjusting for potential confounding factors, there were still significant associations between vaccine uptake and risk perception, knowledge, and attitude. The aOR for risk perception was 1.258 (95% CI: 1.001-1.580), indicating that for each unit increase in risk perception score, the odds of vaccine uptake increased by 1.258 times. The aOR for knowledge was 1.562 (95% CI: 1.154-2.110), indicating that for each unit increase in knowledge score, the odds of vaccine uptake increased by 1.562 times. The aOR for attitude was 1.232 (95% CI: 1.095-1.385), indicating that for each unit increase in attitude score, the odds of vaccine uptake increased by 1.232 times. These findings suggest that individuals with higher levels of risk perception, knowledge, and positive attitude towards the vaccine are more likely to get vaccinated.

### Impact of mandatory Vaccination on Second Dose Uptake of COVID-19 Vaccine

We investigated the impact of forced vaccination on the uptake of the second dose of the COVID-19 vaccine. The results showed that among those who were not forced to vaccinate for the first dose, 108 out of 118 (91.5%) received the second dose, while only 33 out of 45 (73.3%) of those who were forced to vaccinate for the first dose received the second dose. The difference in the uptake of the second dose between the two groups was statistically significant (p = 0.002), and the odds of receiving the second dose were 75% lower (OR: 0.255, 95% CI: 0.101-0.642) in the group that was forced to vaccinate for the first dose compared to the group that was not forced to vaccinate. These findings suggest that mandatory vaccination policies may have a negative impact on the uptake of subsequent vaccine doses.

## Discussion

Vaccines have been crucial in preventing the spread of infectious diseases for decades. With the emergence of COVID-19, vaccination has become more important than ever before. The aims of this study were to investigate COVID-19 vaccine uptake and associated factors, with a focus on the impact of forced vaccination policies on second dose uptake.

The study found that (70.4%) of the residents in Basmaia City, a small city in Baghdad, Iraq, had received at least one dose of the COVID-19 vaccine. This rate is higher than the official percentage of Iraq (46.6%) but lower than the rates reported in developed countries like China (91%) and Canada (90%).(15) Additionally, the study’s finding is consistent with a previous study conducted at a university in Dohuk, Iraq, which reported a vaccine coverage rate of 70.1% among students and staff.(35)

The relatively high vaccine uptake in Basmaia city is encouraging, but it is essential to ensure that individuals who have received the first dose complete the full vaccination course to achieve maximum protection against COVID-19. In this study the majority (86.4%) completed the full vaccination course by receiving the second dose. These results are promising and suggest that efforts to promote vaccine uptake in Basmaia city have been successful.

Before the vaccine campaign was initiated in Iraq, a cross-sectional study was conducted to evaluate the intentions of the population towards the COVID-19 vaccine. The study found that only 56.2% of the participants intended to get vaccinated, while the remainder were hesitant.(36) However, contrary to the well-known intention-behavior gap,(37) the COVID-19 vaccine uptake exceeded the intention rate. This could be attributed to several factors, such as the fear of severe consequences of the disease, community pressure to resume normal activities and overcome the pandemic’s impact on the economy and education, and the expected restrictions on travel and movement for unvaccinated individuals.

The study revealed that the most common reason for getting vaccinated was protection from the disease, indicating that individuals in Basmaia city were aware of the importance of vaccination in protecting themselves against COVID-19. However, a significant proportion of individuals reported being forced to vaccinate, indicating the role of external pressure in increasing vaccine uptake. In contrast, fear of side effects and the perception of not needing the vaccine were the most common reasons for refusing the vaccine. These findings highlight the importance of addressing concerns and providing education to individuals to increase vaccine uptake.

The study identified several factors that were significantly associated with COVID-19 vaccine uptake. These factors included age, sex, job title, education level, perceived risk of COVID-19 transmission, knowledge, and attitude towards the COVID-19 vaccine.

The study results indicated that age was a significant predictor of COVID-19 vaccine uptake, with older individuals being more likely to get vaccinated. This finding is consistent with previous studies,(38)(39) although some studies have shown the opposite.(40) It could be attributed to the fact that older people may have a higher understanding of COVID-19 risk and the benefits of vaccination. And Older adults are at higher risk of severe illness and death from COVID-19, so they may be more motivated to get vaccinated to protect themselves from the disease. However, the finding could also be influenced by Iraq’s vaccination delivery strategy, which gave priority to elderly individuals during the first three months of the vaccination program.

The study found that male Iraqis had a higher vaccination uptake than females, which is consistent with previous reports in Iraq and other countries.(36)(42)(43) However, there is evidence of greater vaccine uptake in females than males in the USA.(44) In Iraq, cultural factors and gender norms may play a role in lower vaccine uptake among females, as they may be more likely to stay at home and have limited access to healthcare and information. Addressing these barriers is crucial to improving vaccination rates and protecting public health. Further research is needed to better understand the factors contributing to low vaccine uptake among women in Iraq and to develop targeted interventions and messaging to promote vaccination.

The finding that participants with a higher education level were more likely to receive the COVID-19 vaccine is consistent with many previous studies.(45)(46)(47) Education is often associated with higher health literacy, which may lead to better understanding of the benefits and risks of vaccination. In addition, individuals with higher education levels may have more access to information and resources related to vaccination, which can increase their vaccine uptake. However, a negative relationship with education level was observed in studies conducted in Kuwait.(48)

The finding that healthcare workers had a 100% vaccine uptake rate is consistent with other studies and can be attributed to their direct exposure to the virus and their understanding of the benefits of vaccination. However, the higher likelihood of vaccine uptake among participants working outside the health field compared to those who were not employed or worked as housewives or retirees suggests that occupation may indeed play a role in vaccine acceptance. This may be due to workplace policies mandating vaccination or providing opportunities for vaccination. On the other hand, retirees and housewives may have less interaction with the healthcare system and fewer opportunities to receive information about vaccines. A study conducted in USA found similar results, with employed individuals having higher vaccine uptake compared to unemployed individuals.(49) Overall, occupational status should be considered when developing strategies to improve vaccine acceptance and uptake among different populations.

The finding that risk perception was significantly associated with vaccine uptake suggests that individuals who perceive COVID-19 as a significant risk are more likely to take actions to protect themselves, including getting vaccinated. There have been several previous studies that have also found a positive association between risk perception and COVID-19 vaccine uptake.(49)(50)(51) This finding is consistent with previous studies that have found a positive association between risk perception and vaccine uptake for other infectious diseases, such as influenza.(52)

It is important to note that risk perception is a complex construct that can be influenced by a variety of factors, including media coverage, personal experiences, and cultural beliefs. Therefore, interventions aimed at increasing vaccine uptake should take into consideration the various factors that influence risk perception.

The findings that knowledge and positive attitude towards the COVID-19 vaccine are significant predictors of vaccine uptake. This is consistent with previous research that has found similar associations between knowledge, attitude, and vaccine uptake in various populations.(53)(54)(55)(56)

One possible explanation for these findings is that individuals who have a better understanding of the vaccine’s benefits, side effects, and effectiveness are more likely to trust and accept the vaccine. Moreover, individuals who have a positive attitude towards vaccines in general may be more inclined to seek out and receive the COVID-19 vaccine.

The results of this study suggest that forced vaccination policies may have a negative impact on the uptake of subsequent vaccine doses.

Specifically, the study found that individuals who were not forced to vaccinate for the first dose had a significantly higher uptake of the second dose compared to those who were forced to vaccinate for the first dose.

This finding is consistent with previous studies that have investigated the impact of mandatory vaccination policies on vaccine uptake. For example, a study conducted in Italy found that the introduction of mandatory vaccination policies for children led to a decrease in vaccination coverage.(57)Similarly, a study conducted in France found that mandatory vaccination policies for healthcare workers led to a decrease in vaccine uptake.(58)

There are several potential reasons why forced vaccination policies may have a negative impact on vaccine uptake. One possibility is that these policies may lead to feelings of resentment or mistrust towards the government or healthcare system. Additionally, some individuals may view mandatory vaccination as a violation of their personal autonomy or beliefs.

To address this issue, it may be beneficial to focus on education and communication efforts to increase vaccine uptake rather than resorting to mandatory vaccination policies.

### Limitation

One of the limitations of our study is that it was cross-sectional, which means that the data was collected at a specific point in time. As a result, the study only provides a snapshot of the community’s response to COVID-19 vaccination at that particular time.

Secondly, since the study relied on self-reported data, there is a possibility of reporting bias, where participants may have answered questions based on what they thought was expected of them, rather than their actual beliefs or behaviors.

Finally, one possible limitation is that participants who declined to enroll may have done so because they did not want to admit to certain behaviors or beliefs that they perceived as socially undesirable. This could lead to underrepresentation of certain groups in the study population.

## Conclusion

In conclusion, the prevalence of COVID-19 vaccine uptake among the general population in Basmaia City, Baghdad was found to be high at 70.4%, which is higher than the officially reported rates. with the majority of those vaccinated having received both doses. The most common reason for getting vaccinated was for protection against the disease, while fear of side effects and not seeing a need for the vaccine were the most common reasons for refusing it. Factors associated with vaccine uptake included being male, older, having a higher level of education, being employed (especially in healthcare), having a higher risk perception, and having a positive attitude and knowledge about the vaccine. Additionally, our study found an association between mandatory vaccination and higher rates of second dose uptake.

### Recommendation

Based on the findings of this study, several recommendations can be made to increase COVID-19 vaccine uptake in Basmaia City, Baghdad. First, targeted educational campaigns should be developed to address the low vaccine uptake among younger age groups, particularly females. These campaigns should focus on increasing knowledge about the vaccine, its benefits, and potential side effects.

Second, on-site vaccination options should be made available to increase vaccine uptake among unemployed individuals and those who are homebound due to illness or disability. This could involve providing mobile vaccination clinics or allowing individuals to receive the vaccine at home.

Third, the mandatory vaccination policy should be reviewed to include rules that mandate the uptake of the second dose of the vaccine. This would ensure that individuals receive the full benefit of the vaccine and reduce the risk of breakthrough infections.

Finally, further studies are needed to better understand the reasons behind vaccine hesitancy and refusal in this population, and research on the effectiveness of targeted interventions, such as community outreach programs and education campaigns, in increasing vaccine uptake among different subgroups of the population.

### Dedication

To my beloved wife, parents, and daughters, I dedicate this study with utmost gratitude and admiration. Above all, I want to express my profound gratitude to God for guiding me through this academic journey. Your unwavering support, patience, and love have been my constant motivation, and I couldn’t have achieved this without you. Your trust and belief in me have been my anchor, providing a sense of purpose and direction. This accomplishment is not just mine, but it belongs to all of us. Thank you for being my guiding light and inspiration.

## Data Availability

All data produced in the present study are available upon reasonable request to the authors

## Acknowledgments

I would like to express my deepest appreciation to my supervisor for their valuable guidance, encouragement, and support throughout this research project. I am also grateful to the management of Basmaia City for granting me access to the necessary resources and facilities to complete this study. Without their assistance, this research would not have been possible. Also I would like to acknowledge the contributions of all the study participants who willingly gave their time and shared their experiences to make this project a success. Lastly, I would like to acknowledge the contribution of the AI language model (chatGPT), which has assisted me in the writing and editing process.

